# The MAGMA pipeline for comprehensive genomic analyses of clinical *Mycobacterium tuberculosis* samples

**DOI:** 10.1101/2023.10.04.23296533

**Authors:** Tim H. Heupink, Lennert Verboven, Abhinav Sharma, Vincent Rennie, Miguel de Diego Fuertes, Robin M. Warren, Annelies Van Rie

## Abstract

**Background:** Whole genome sequencing (WGS) holds great potential for the management and control of tuberculosis. Accurate analysis of samples with low mycobacterial burden, which are characterized by low (<20x) coverage and high (>40%) levels of contamination, is challenging. We created the MAGMA (Maximum Accessible Genome for *Mtb* Analysis) bioinformatics pipeline for analysis of clinical *Mtb* samples.

**Methods and results:** High accuracy variant calling is achieved by using a long seedlength during read mapping to filter out contaminants, variant quality score recalibration with machine learning to identify genuine genomic variants, and joint variant calling for low *Mtb* coverage genomes. MAGMA automatically generates a standardized and comprehensive output of drug resistance information and resistance classification based on the WHO catalogue of *Mtb* mutations. MAGMA automatically generates phylogenetic trees with drug resistance annotations and trees that visualize the presence of clusters. Drug resistance and phylogeny outputs from sequencing data of 79 primary liquid cultures were compared between the MAGMA and MTBseq pipelines. The MTBseq pipeline reported only a proportion of the variants in candidate drug resistance genes that were reported by MAGMA. Notable differences were in structural variants, variants in highly conserved *rrs* and *rrl* genes, and variants in candidate resistance genes for bedaquiline, clofazmine, and delamanid. Phylogeny results were similar between pipelines but only MAGMA visualized clusters.

**Conclusion:** The MAGMA pipeline could facilitate the integration of WGS into clinical care as it generates clinically relevant data on drug resistance and phylogeny in an automated, standardized, and reproducible manner.

**Key points:**

- Accurate analysis of clinical samples is challenging when samples have high levels of contamination and low *Mycobacterium tuberculosis* genome coverage
- When analyzing primary liquid (MGIT) cultures, the MAGMA pipeline generates clinically relevant drug resistance information (including major, minor and structural variants) and phylogeny in an automated, standardized and reproducible way.
- MAGMA-generated phylogenetic trees are annotated with drug resistance information and updated with every run so that they can be used to make clinical or public health decisions
- MAGMA reports drug resistance variants for all tier 1 and tier 2 candidate drug resistance conferring genes, with interpretation of their relevance to drug resistance (associated with drug resistance, not associated with drug resistance or unknown significance) based on the WHO catalogue of mutations in *Mycobacterium tuberculosis*.

## Background

Genomic data from *Mycobacterium tuberculosis* (*Mtb)* can provide clinically relevant insights such as the diagnosis of drug resistance and the identification of transmission clusters [1]. Whole genome and targeted sequencing of *Mtb* have higher discriminatory power than the conventional spoligotyping and MIRU-VNTR genetic typing methods [2]. Next generation sequencing is also superior to the current combination of line probe assays and phenotypic drug sensitivity tests used in routine diagnostic laboratories, as sequencing can comprehensively define the drug resistance profile in a single assay.

Sequencing *Mtb* from sputum or primary liquid culture isolates could greatly contribute to tuberculosis management as results can be used to inform clinical or public health decisions [3]. Analysis DNA extracted directly from sputum samples is also important as it avoids culture bias and thus better reflects in-patient *Mtb* population diversity. Although sequencing directly from sputum is already possible, whole genome sequencing (WGS) of *Mtb* is currently mainly performed on DNA extracted after one or more culturing steps to analyze purified *Mtb* genomic data. WGS analysis directly from sputum is much more challenging due to the high levels of non-*Mtb* contaminants and low genomic coverage of *Mtb* [4]. Simulation studies showed that commonly used *Mtb*-specific pipelines (MTBseq and UVP) can experience difficulties when contaminant levels exceed 40% or when depth of coverage is below 25x [5, 6]. Recent approaches have aimed to circumvent this by enriching *Mtb* DNA *in vitro* or *in silico*. *In vitro* methods include enzymatic removal of non-*Mtb* contaminants [7], *Mtb*-specific DNA-capture probes [8, 9] or multiplex-amplification of drug resistance-conferring regions [10, 11]. *Mtb* enrichment *in silico* can be achieved by classifying reads and rejecting any reads not classified as *Mtb* [12]. These enrichment methods are not yet used in clinical practice as they are expensive, time-consuming, require additional computing infrastructure, and can produce bias for the probed regions or for regions of the genome where reads are more readily mapped.

In recent years, improvement in the analysis of complex sequencing data, particularly with respect to variant calling and filtering [13, 14], have been extensively implemented for the analysis of human genomes but have seen limited applications for the analysis of bacterial genomes. We developed MAGMA (Maximum Accessible Genome for *Mtb* Analysis) pipeline, a novel bioinformatics pipeline by implementing the ‘compleX Bacterial Sample’ XBS variant calling core [6]. The XBS variant calling core is a bioinformatics workflow, in the form of a set of scripts, that can be implemented at the core of a pipeline to obtain accurate bacterial genomic variants from complex biological samples. The XBS variant calling core relies on two principles (1) joint variant calling, where variants are called for a cohort rather than a single sample, enabling confident variant calling in low coverage genomes and (2) Variant Quality Score Recalibration, where machine learning is used to identify genuine genomic variants. The latter filters out false positive genomic variants introduced by contaminants and uses the model for genuine genomic variants to identify true positive genomic variants in complex genomic regions [6]. This process makes it possible to analyse additional SNPs from regions that are normally discarded, thereby significantly extending the accessible genome for *Mtb* given the prevalence of complex regions in the *Mtb* genome [15].

In the method section of this manuscript we describe the overall architecture of the MAGMA pipeline, including the key design choices made to address critical issues. In the results section we present the drug resistance and phylogeny outputs for 79 primary liquid cultures and compare the drug resistance and phylogeny outputs generated by MAGMA to those generated by MTBseq, a commonly used open-source *Mtb* pipeline.

## Results

To show the functionalities of the MAGMA pipeline, we present the analysis of sequencing data obtained from primary cultures from 79 consecutive participants in the SMARTT trial (Sequencing Mycobacteria and Algorithm-determined Resistant Tuberculosis trial, *Clinicaltrials.gov Identifier NCT05017324*) [3]. For this trial, primary liquid cultures were processed in real time in batches of 4-6 samples on an Illumina MiniSeq for individualized WGS-guided treatment decision-making.

### Computational Runtime and resource usage

The analysis of 79 samples using AWS batch took 4h to complete and required a total of 204.9 CPU hours. The size of the input dataset directory was 47.5 GB, the results directory was 0.29 GB and the work directory was 418.9 GB, corresponding to a total data footprint of the analysis of 466.7 GB.

To determine the computational runtime and resource usage when MAGMA is used for clinical decision-making in a low-resource setting, we also documented the analysis of batch of 4 samples, together with a reference set of 334 previously processed reference samples, run on a 2011 Linux laptop with 8 GB of RAM and 8 CPU cores and an Intel® Core^TM^ i7-2630QM CPU (2 GHz). The analysis required 4h 52m to complete with a cap of 6GB RAM use. A total of 28.9 CPU hours was required for the analysis. The size of the input dataset directory was 2 GB, and the final directory size including the results and intermediate directories was 7.8 GB.

### Quality Check and Mapping outputs

The base quality of the 79 samples was high, with an average Phred quality score of 35.6 (range 33.8 to 36.5) (Figure 1A, supplementary table 1). Most samples had a poly-modal guanine-cytosine (GC) content distributions, with the most prominent peaks matching the 65.6% GC content of the H37Rv reference genome. Four samples (MM-054, LP-239, XP-244, LP-027, LP-321) displayed a higher than expected GC content (Figure 1B) [16].

**Figure 1:**
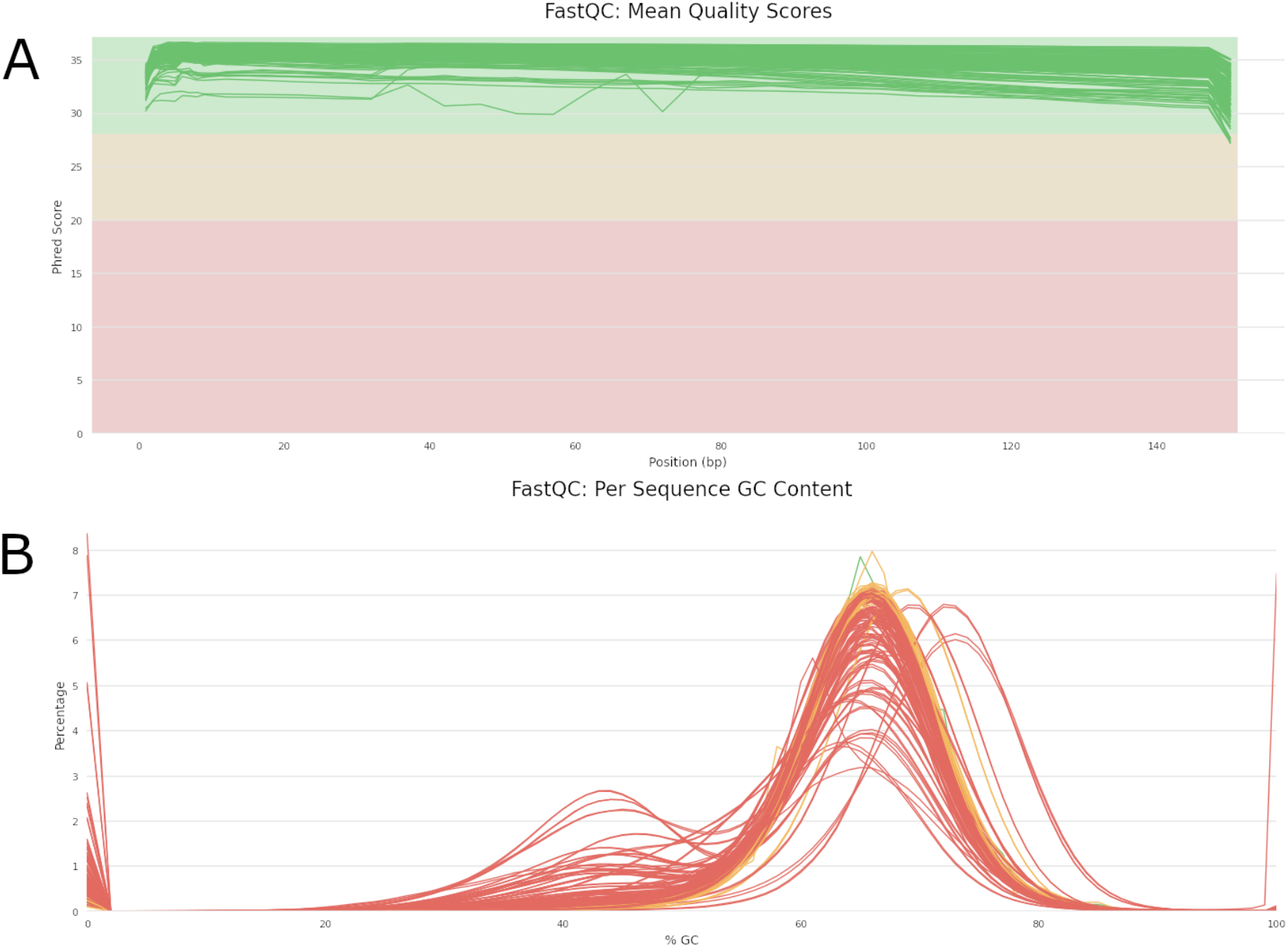
MultiQC visualization of per-base pair quality (A) and percentage Guanine-Cytosine (%GC) content curves (B) of raw sequencing fastQ files obtained from DNA extracts from 79 primary liquid cultures.

Mapping percentage (average 58.0, range 0.03%-87.5%) and depth of coverage (average 107x range 0x-308x) varied by sample. Eight samples were automatically excluded by the MAGMA pipeline. Three because the major strain did not exist at a frequency exceeding 0.80 (LP-022, LP-250, MM-054), possibly indicating the presence of mixed infection and five samples (FD-047, LP-027, LP-239, LP-321, XP-244) because of insufficient depth (>10x required) and/or breadth of coverage (>90% required). The remaining 71 samples passed quality control and were included in subsequent analyses.

### Variant Calling

In the best performing VQSR filtering run used for downstream analysis, none of the resulting 9202 filtered variants had a negative VQSLOD score when applying the 99.9% sensitivity threshold. A 100% sensitivity threshold reported an additional 100 variants of which 83 variants with a negative VQSLOD score, corresponding to the presence of 0.89% of likely false positives variants in the unfiltered dataset (Figure 2).

**Figure 2:**
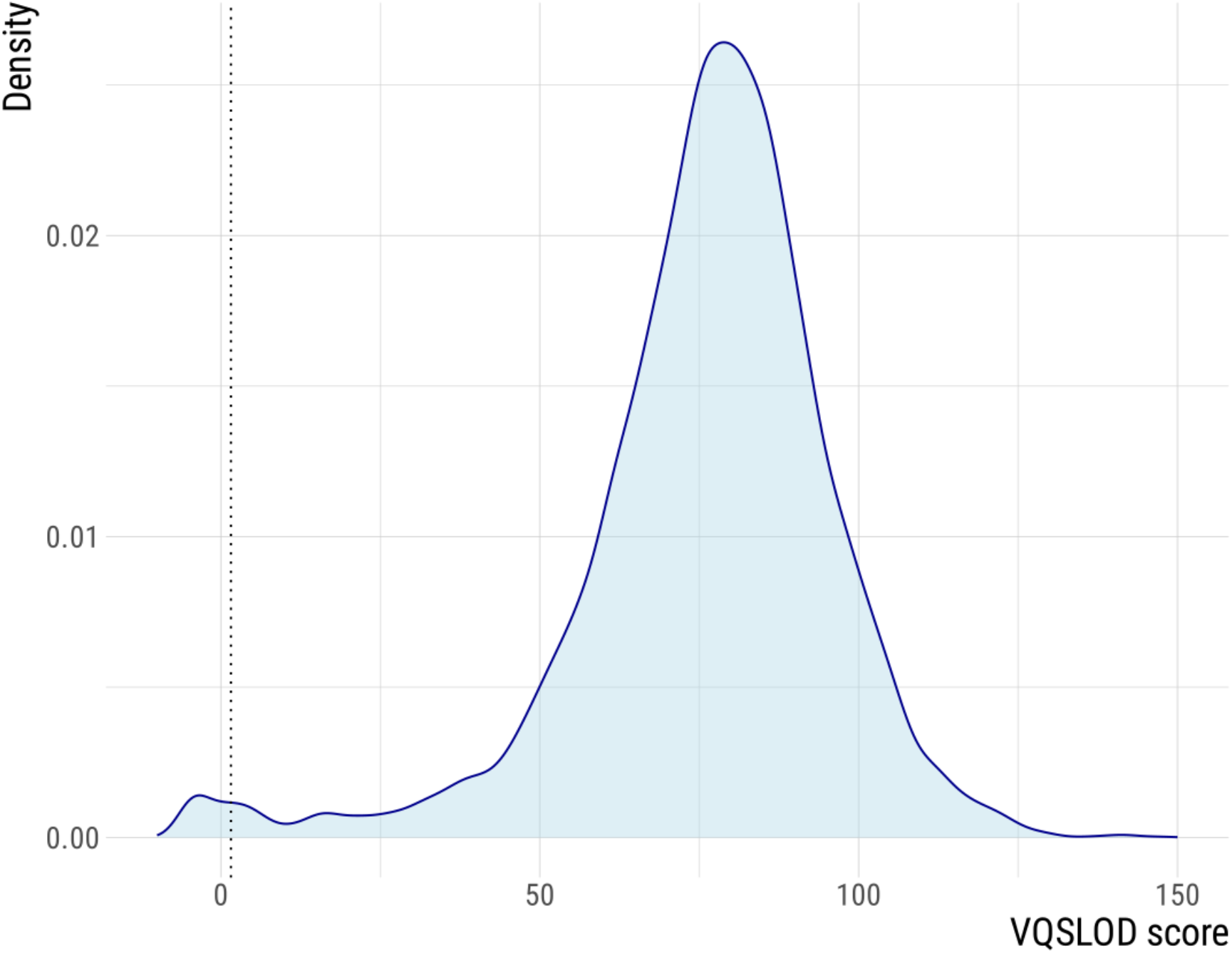
Density distribution of the variant quality score log-odds (VQSLOD) for the identified SNP variants. Variants with scores below zero are most likely false positives according to the Variant Quality Score Recalibration (VQSR) models, a 100% sensitivity would include these variants. MAGMA uses the 99.9% sensitivity cut-off for filtering, indicated by the dashed line, thereby excluding variants with negative VQSLOD scores.

### Drug resistance output

The MAGMA pipeline outputs comprehensive drug resistance information for 23 anti-TB drugs for each sample in a per-sample excel table (supplementary table 2). The resistance classification for each drug is based on classification in the WHO catalogue. In addition, the excel file lists the classification of the type of variant (e.g. upstream gene variant), the method by which it was detected (XBS, LoFreq, or delly), the frequency at which the variant was detected, whether the variant is listed in the WHO catalogue, the source annotation within the WHO catalogue as well as a direct html link to the WHO catalogue. For variants classified as ‘of unknown significance’ in the WHO catalogue, the excel file lists in how many phenotypically resistant and sensitive samples the variant was observed and shows the fraction. In the “Source” column notes variants located within a Tier 1 or Tier 2 gene not listed in the WHO catalogue.

### Phylogenetic output

The MAGMA pipeline outputs two phylogenetic trees. Both phylogenetic inferences exclude regions associated with drug resistance, minor variants, structural variants, insertions and deletions. The default tree includes complex *Mtb* genomic regions (IncComplex), the other tree (ExComplex) excludes them (supplementary figures 1A and 1B). MAGMA also produces text files which contain lineage information and drug resistance profiles for each sample. The phylogenetic tree (treefile) and the associated text files can be uploaded to iTol (https://itol.embl.de) to generate annotated phylogenetic trees (Figure 3). For each of the phylogenetic trees (IncComplex and ExComplex), two figtree files are generated to visualize the presence of clusters, one using a 5 SNP and another using a 12 SNP cut-off. When imported into the FigTree desktop application, these figtree files are automatically annotated so that the tip label of samples belonging to a cluster have the same color (Figure 4).

**Figure 3:**
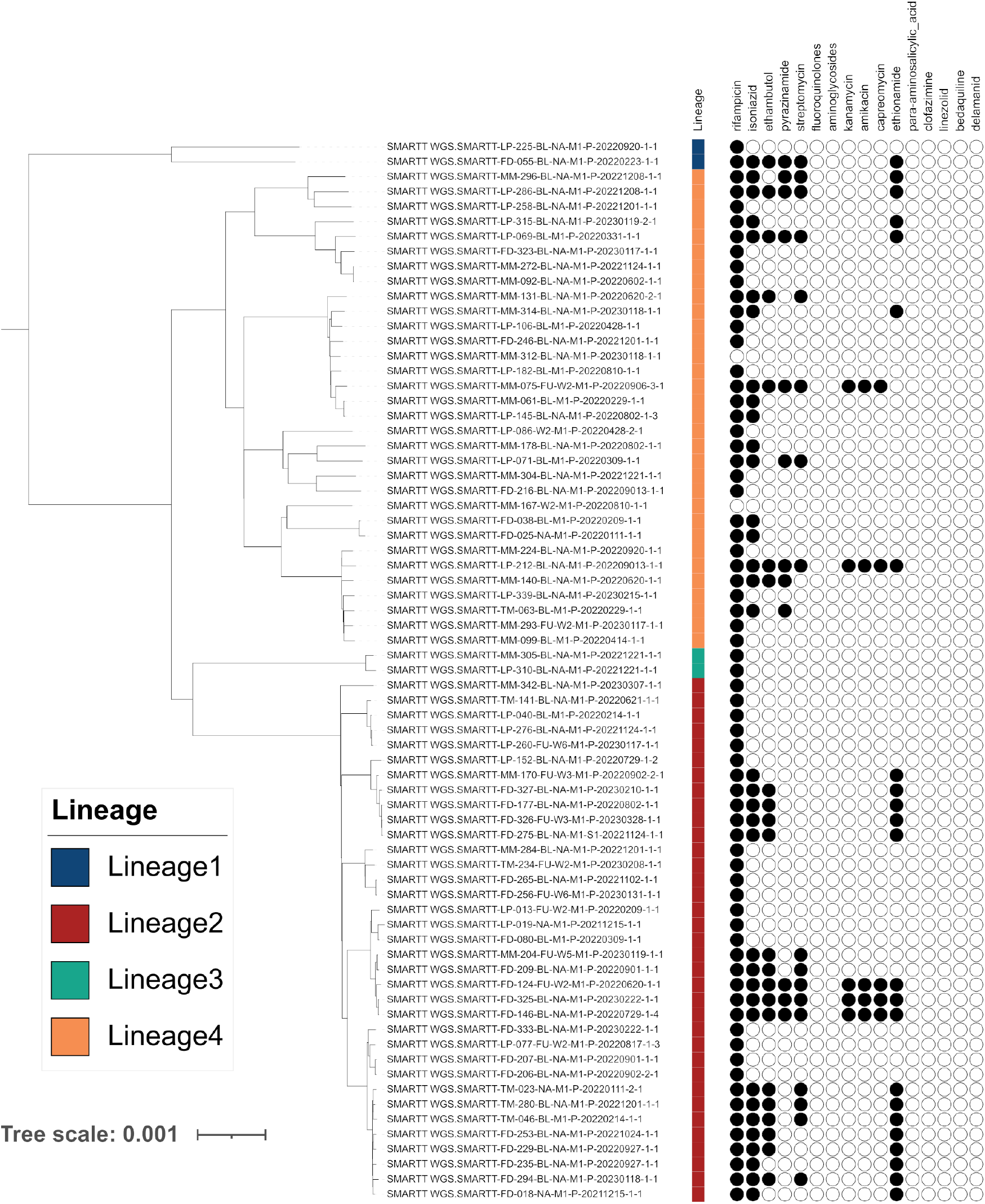
iTOL visualizations of the MAGMA ExComplex phylogeny annotated with drug resistance and lineage, for 71 primary liquid culture samples that passed MAGMA quality control

**Figure 4:**
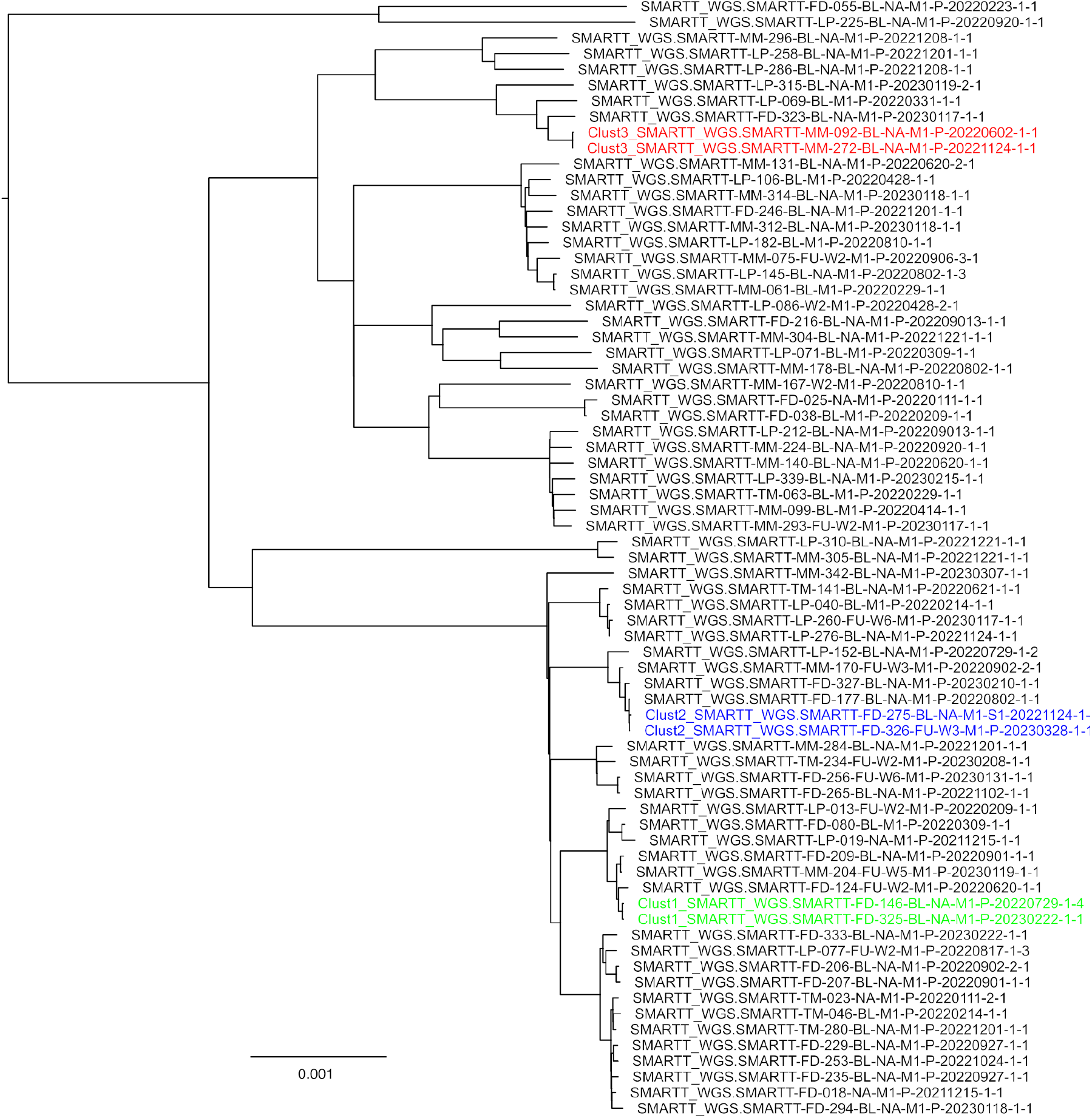
FigTree visualization of the phylogeny annotated with 5SNP cluster information (each color represents a different cluster) and indicating the newly added samples to the phylogenetic tree

### Comparison of drug resistance output: MAGMA and MTBseq

To demonstrate the differences between the MAGMA and MTBSeq pipeline, we compared in detail the output generated for the primary liquid culture collected at the start of treatment in one patient (TM280, lineage 2.2.1, median coverage 163, breath of coverage 98.6) (Supplementary tables 2 for MAGMA and 3 for MTBseq) The drug resistance information generated by the MAGMA and MTBseq pipelines for patient TM280 was compared for all variants identified in tier1 and 2 genes for the 15 drugs included in the 2021 WHO catalogue of mutations in *Mtb* [42] (Table 1).

**Table 1:**
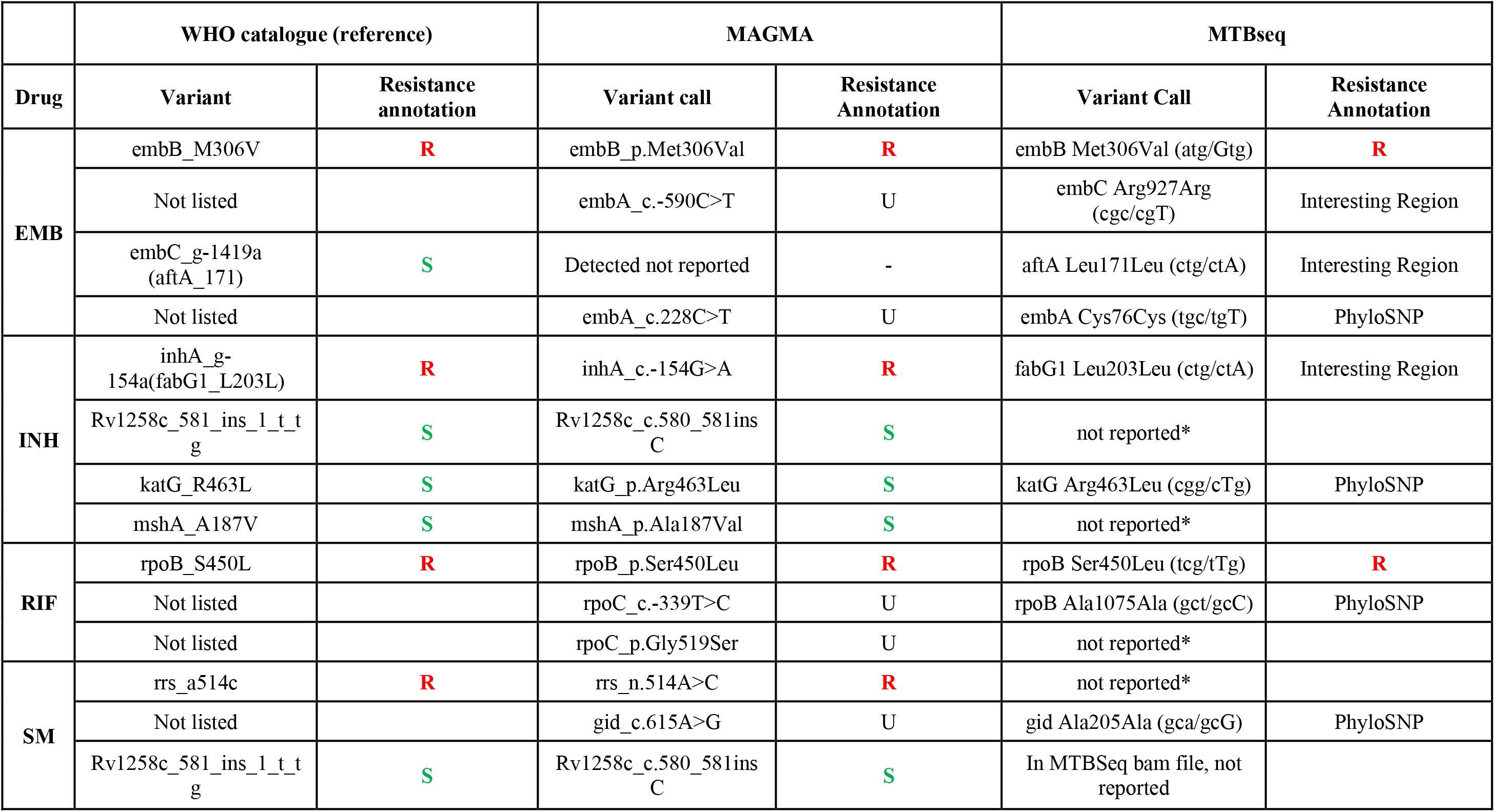
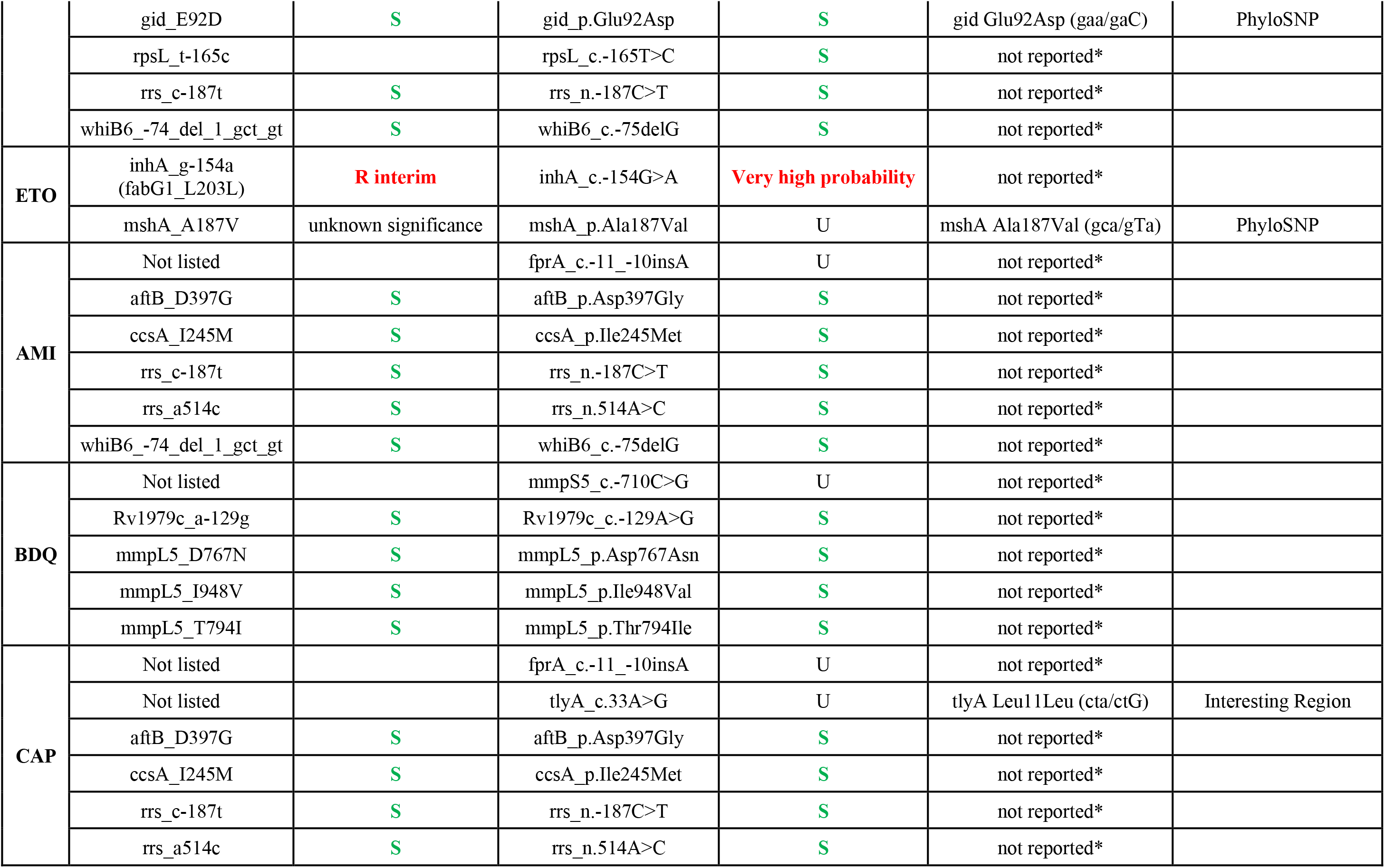
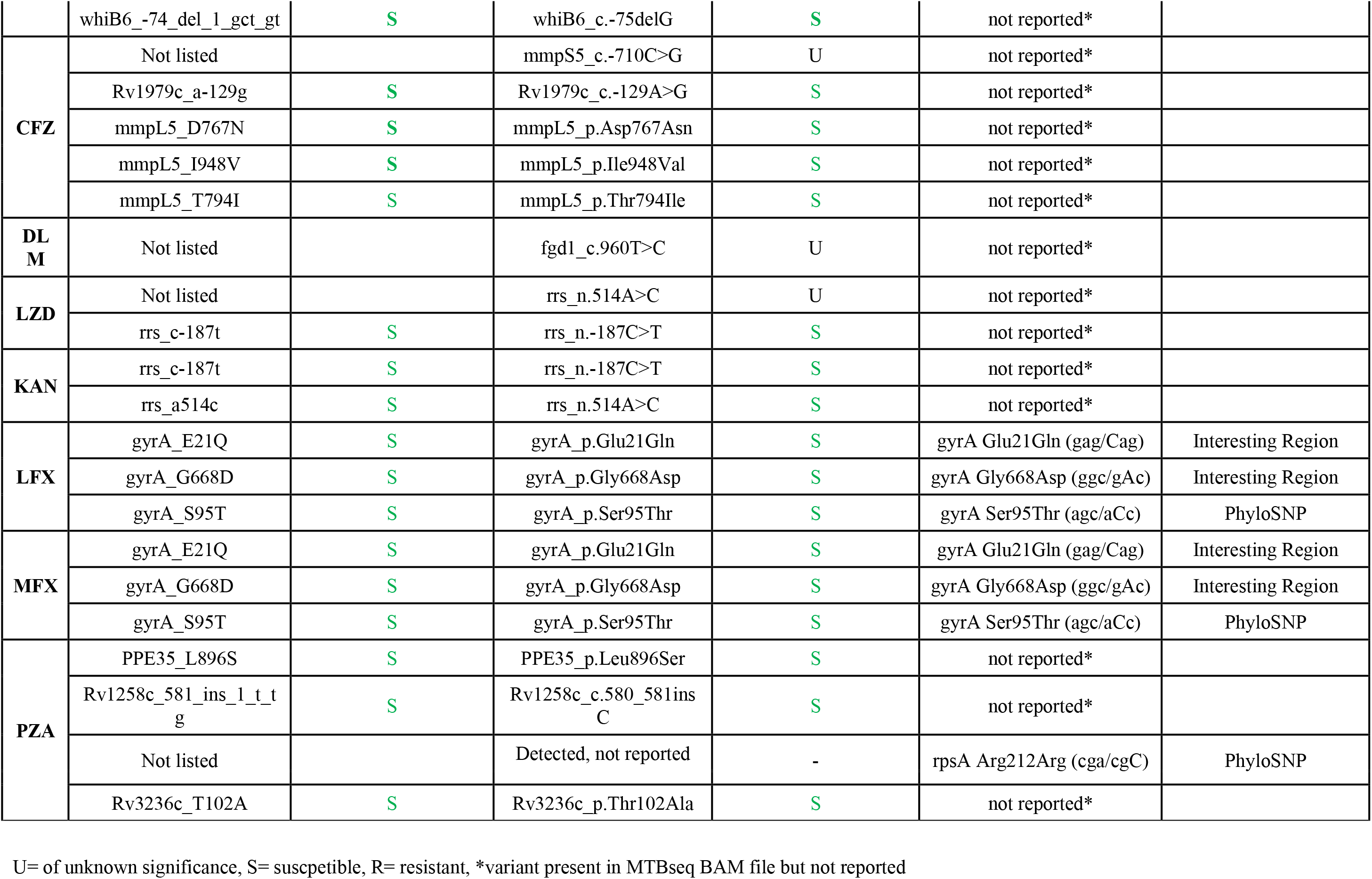
Drug resistance information as reported by MAGMA and MTBseq, including the variant and resistance annotation in the WHO catalogue of mutations in *Mycobacterium tuberculosis*.

**Table 2:** Comparison of resistance reported by MAGMA and MTBSeq pipelines.

|  | Number of drug resistant samples |  | Discordances in drug resistance reporting |  |  |  |  |
| --- | --- | --- | --- | --- | --- | --- | --- |
|  | MAGMA | MTBSeq | N | Variant | WHO catalogue | MAGMA | MTBSeq |
| RIF | 68 | 59 | 1 | rpoB_His455Leu | R | R | Variant not reported* |
|  |  |  | 1 | rpoB_His455Leu | R | R | U<br>(Interesting_Region) |
|  |  |  | 6 | rpoB_Ser450Leu | R | R | Variant not reported* |
|  |  |  | 1 | rpoB_Leu430Pro | R | R | Variant not reported* |
| INH | 35 | 27 | 9 | inhA_-154G>A | R | R | U<br>(Interesting_Region) |
|  |  |  | 1 | full katG deletion | not listed | U | R**<br>katG_Leu141Phe |
| PZA | 10 | 15 | 3 | pncA_His57Pro | U | U | R |
|  |  |  | 1 | pncA_His57Gln | U | U | R |
|  |  |  | 1 | pncA_Leu35Arg | not associated with<br>R-interim | very low prob-<br>ability of R | R |
| EMB | 22 | 24 | 1 | embA_-11C>A | U | U | R |
|  |  |  | 1 | embA_-16C>G | U | U | R |
| ETO | 12 | 15 | 3 | ethA_Tyr276His | U | U | R |
| AMI | 5 | 4 | 1 | rrs_1401A>G | R | R | Variant not reported* |
| CAP | 5 | 4 | 1 | rrs_1401A>G | R | R | Variant not reported* |
| **KAN** | 5 | 4 | 1 | rrs_1401A>G | **R** | **R** | Variant not reported* |
| **STM** | 16 | 11 | 3 | rrs_514A>C | **R** | **R** | Variant not reported* |
|  |  |  | 1 | rrs_517C>T | **R** | **R** | Variant not reported* |
|  |  |  | 1 | rpsL_Lys43Arg | **R** | **R** | Variant not reported* |
| **BDQ** | 0 | 0 | 0 |  |  |  |  |
| **CFZ** | 0 | 0 | 0 |  |  |  |  |
| **DLM** | 0 | 0 | 0 |  |  |  |  |
| **LZD** | 0 | 0 | 0 |  |  |  |  |
| **LFX** | 9 | 9 | 0 |  |  |  |  |
| **MFx** | 9 | 9 | 0 |  |  |  |  |

Of the 32 unique variants reported by MAGMA, nine were not listed in the WHO catalogue and 16 were not reported by MTBseq. Of the 16 unique variants reported by MTBseq, five were not listed in the WHO catalogue and two were not reported by MAGMA. The variant *embC*_g-1419a (*aftA*_171) was detected by MAGMA but not reported because *aftA* is not a tier 1 or 2 gene and it lies more than 200 bp upstream of the *embC* gene. Susceptible variants are only reported when they fall no more than 200 bp up- and downstream of the furthest resistance conferring variant of any tier 1 or 2 gene. The *rpsA* Arg212Arg variant was not reported because the rpsA gene is not a tier 1 or 2 gene. In total, 14 variants were reported by both pipelines, sometimes using different annotations, for example *inhA*_c.-154G>A in MAGMA and *fabG1*_Leu203Leu (ctg/ctA) in MTBseq. When variants were detected by both pipelines, they were not always reported for the same drugs. For example *mshA*_p.Ala187Val was only listed for isoniazid (INH) by MTBseq but listed for both and ethionamide (ETO) by MAGMA. The *mshA* gene is included as a tier 2 gene for both ethionamide and isoniazid in the WHO catalogue. With respect to the resistance annotation for reported variants, MAGMA classified five variants as resistant conferring, corresponding to the WHO classification, whereas only two variants were reported as resistance conferring by MTBseq. This resulted in a different resistance profile output by the two pipelines: resistance to INH, RIF, EMB, ETO, and SM by MAGMA versus resistance to EMB and RIF by MTBseq. For the 15 drugs, 38 variants were classified as not associated with resistance by the WHO catalogue and susceptible by MAGMA. Nine variants were classified as a PhyloSNP by MTBseq, suggesting a susceptible phenotype. In addition, 14 variants were classified as of unknown significance by MAGMA and 8 variants were classified as located in an interesting region by MTBseq.

Table 2 compares the number of drugs classified as resistant in the 79 patients, limited to the 15 drugs listed in the WHO catalogue. In total, 170 identical resistance calls were made for both pipelines and 37 calls differed between the two pipelines. Differences between MTBseq and MAGMA calls were due to variants not being reported by MTBseq, variants reported as located in an interesting region by MTBseq while listed as “associated with resistance” in the WHO catalogue, or variants listed as resistance conferring by MTBseq while listed as “of unknown significance” in the WHO catalogue. MAGMA also detected a 20 kb deletion including the full *katG* gene that was not detected by MTBseq. The coverage of this region in MTBseq was lower than in neighboring regions, indicating that the *katG* gene indeed does not exist in this sample. MTBseq maps reads on the deleted region, some of which contain variant *katG*_p.Leu141Phe, which MTBseq then reported as resistance-conferring.

### Comparison of phylogenetic output: MAGMA and MTBseq

MAGMA excluded eight samples because of low quality, resulting in a phylogenetic tree for 71 samples (supplementary figure 2B). Using default settings, MTBseq generated a tree for all 78 samples (supplementary figure 2A). When applying coverage-based filters, as recommended in the MTBseq repository (https://github.com/ngs-fzb/MTBseq_source), the same eight low-quality samples were manually excluded, resulting a similar phylogenetic tree compared to the ExComplex MAGMA tree (supplementary figure 3). Heatmaps show that the default MTBseq tree has poor structure without clear lineation between major *Mtb* lineages, while the MAGMA and filtered MTBseq tree have the correct tree structure (supplementary figure 4). MAGMA identified a total of 8514 non-DR variant positions (7184 for the phylogenetic comparison without complex regions), MTBseq 7296 non-DR variants when excluding the eight low-quality samples.

## Discussion

WGS is increasingly used in high income countries for the management of TB but is not yet implemented in high TB burden settings, in part because the required computing infrastructure and bioinformatics expertise is lacking. To help overcome this challenge, we developed MAGMA, an automated comprehensive bioinformatics pipeline specifically created for the analysis of WGS data obtained from clinical samples.

Analysis of 79 primary clinical liquid cultures demonstrated that both the MAGMA and MTBSeq pipelines can analyze data with variable *Mtb* DNA to total DNA mycobacterial load and large amounts of contaminating sequences. MAGMA was more comprehensive in defining drug resistance as it detects major variants, minor and structural variants in all tier 1 and tier 2 candidate drug resistance genes for all 23 TB drugs. In contrast, MTBseq only identifies major variants defined as present at a >75% frequency. Furthermore, as predicted by a simulation study [6], MAGMA has higher accuracy than MTBseq and identifies more variants in clinical samples due to different approaches to variants filtering. MTBseq employs hard filters where each variant needs to be covered by ≥4 forward and reverse reads, ≥ 4 calls with ≥20 Phred score, and ≥ 75% frequency, where contaminants may result in false positive forward and reverse read counts or negatively impact the frequency threshold. In contrast, MAGMA removes contaminants through strict mapping and machine-learning based variant filtering (VQSR) and increases the variant calling accuracy for samples with low depth of genome coverage by using joint variant calling. The difference in accuracy was especially notable for structural variants, which are hard to detect by WGS, for variants in in highly conserved regions such as the *rrs* and *rrl* genes, and variants in candidate resistance genes for Bedaquiline, clofazamine and delenamide. MAGMA summarizes the drug resistance information in a comprehensive, standardized output and interprets the variants based on the WHO catalogue as the reference. This results in a clinically relevant drug resistance profile that can be used to guide treatment. In contrast, MTBseq uses a custom classification, resulting in drug resistant classifications that differ from the WHO catalogue.

MAGMA also automatically generates multiple phylogenetic tree including trees including and excluding complex repetitive regions of the Mtb genome, with samples tip-labelled to highlight the new samples added to the database, and a color-coded cluster tree at both 5 and 12 SNP cut-off. The phylogenetic and cluster information can assist public health programs with implementing precision public health by guiding targeted source tracing. After manual filtering by the user, MTBseq also generates a phylogenetic tree excluding complex repetitive regions.

Several innovations allowed the MAGMA pipeline to overcome most of the open issues for *Mtb* WGS identified by Meehan et al. [1] and extends the standards of current *Mtb* pipelines, of which MTBseq is the most commonly used (Table 3). First, by using a long seed length during mapping and VQSR variant filtering, computationally intensive metagenomic analyses is avoided and contaminants are filtered out effectively. Second, use of TBProfiler version 5.0 with the 2021 WHO catalogue of mutations in *Mtb* set as the reference for calling drug resistance variants results in the use of a transparent, comprehensive standardized approach to defining the genomic drug resistance profile of clinical samples. In addition, the MAGMA pipeline lists variants in candidate drug resistance genes (tier 1 and 2) that were not yet included in the 2021 catalogue, reports variants in candidate resistance genes for drugs not listed in the WHO catalogue (such as rifabutin and meropenem), reports minor variants identified by LoFreq to detect the presence of hetero-resistance, and identifies variants in complex genomic regions which could contain novel drug resistance-conferring mutations. Furthermore, the functional annotation of all variants with a prediction of their impact on the protein produced by SnpEff is reported by MAGMA. Third, because the use of any threshold selection for phylogeny can be problematic, MAGMA identifies transmission clusters at both 5 and 12 SNPs, thresholds that can be changed as our knowledge on the most appropriate threshold improves. The MAGMA pipeline identifies all (sub-)lineages that are present at higher levels than the LoFreq significance threshold. Based on the presence of all (sub-) lineages for all samples. Based on this lineage information, samples where there is no majority strain (i.e., minimal presence of 80%), are excluded. Within host microevolution can also be inferred by analysis of samples of the same patient, where microevolution can be inferred by changes in the presence or frequency of minor variant. Fourth, the MAGMA pipeline also pushes the current standards as its XBS core can accurately analyze sequence data from primary cultures. Simulation data suggests that MAGMA can also analyze DNA extracted directly from sputum, but this needs to be further confirmed in clinical studies[6].

**Table 3:**
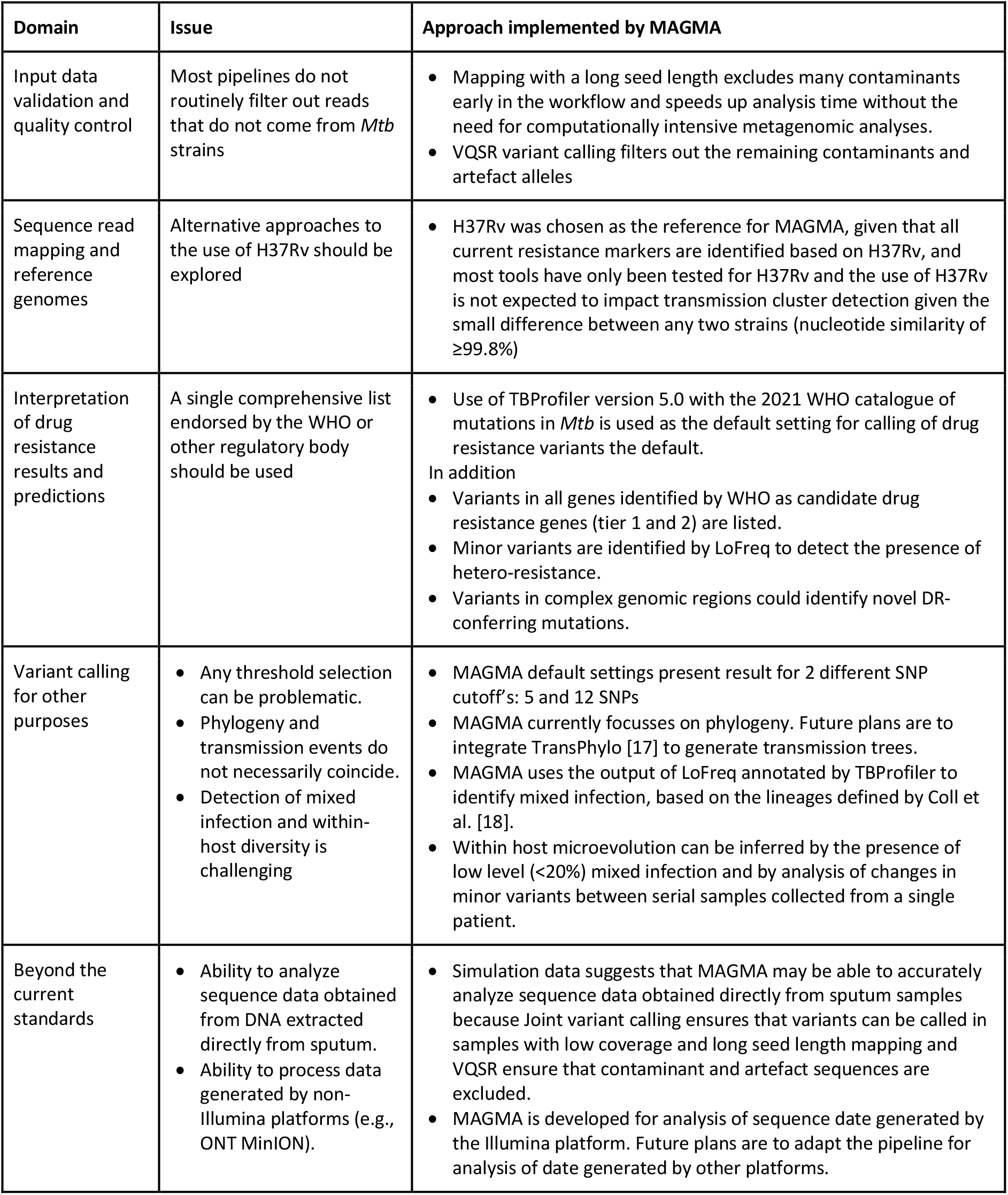
MAGMA approach to overcome open issues in whole genome sequencing of *Mycobacterium tuberculosis* [1].

Another important strength of MAGMA, similar to MTBSeq, is its free online availability and that the analyses can be carried out on a variety of platforms including laptops high performance computing systems, and cloud computing environments. The pipeline is highly automated which allows end-users without a strong background in bioinformatics to use MAGMA to perform WGS analyses of clinical samples. Nevertheless, the interpretation of the resistance profiles and transmission clusters may still be challenging for clinicians, microbiologists in reference laboratories, and public health practitioners. Reducing the proportion of variants classified a ‘of unknown significance’ with regards to drug resistance and WHO endorsement and/or ISO-accreditation of the MAGMA workflow for *Mtb* resistance calling, as for *abritAMR* [19], could aid the implementation and acceptance in clinical use. Integration of TransPhylo [17] to generate transmission trees could facilitate the interpretation of the relevance of the phylogenetic information by public health practitioners. We are therefore working on the development of a web-platform that streamlines the interpretation of the MAGMA outputs for end-users to further improve the user-friendliness of the pipeline.

The MAGMA pipeline also has many technical strengths. First, the use of GATK VSQR joint variant calling, unique in MAGMA as compared to other *Mtb* pipelines, results in a very high sensitivity for variant calling (98.8% in a Sanger sequencing confirmed dataset) and a very high accuracy (F_1_ scores consistently above 0.95 with >20x *Mtb* coverage, even with contamination levels above 80% [6]). In the 79 clinical samples, none of the variants were likely false positive as all had a positive VQSLOD scores meaning that they had a better fit with the “true positive” than the “true negative” model. The high sensitivity does require that minor variants are interpreted with caution. In particular, minor variants called by LoFreq (which does not stringently filter using the VSQR joint filtering) could be contaminant sequences of genomic regions that share high similarity with *Mtb*, such as the *rrs* and *rrl* genes. Interpretation of the clinical relevance of minor variants and variants that have not been statistically associated with resistance or susceptibility will remain challenging and requires expert opinion [20]. Second, the machine-learning based variant filtering (VQSR) is extended across the part of the complex genomic regions, which increases the total genetic resolution by 9% for subsequent analyses [6]. MTBseq and other pipelines remove such variants for phylogenetic inferences and may suggest to also exclude those variants that lie in close proximity of each other (e.g. those in a 12bp window).

The strengths of the MAGMA pipeline should be viewed in the light of some limitations. First, sequencing reads produced by Oxford Nanopore Technology or PacBio platforms cannot yet be processed but this is planned for the near future. Second, VQSR INDEL filtering is not yet implemented because a “truth set” for INDELs is not yet available. While this could lead to false positive INDEL calls, we believe this design choice is correct for the context of clinical care as it aims to avoid missing potentially drug resistance-conferring INDELs. When INDEL “truth datasets” become available, the INDEL VSQR can be easily implemented in the MAGMA pipeline. Third, MAGMA uses the H37Rv reference genome. Consequently, genes that are absent in the reference genome but present in different lineages of *Mtb* are not analyzed, an effect that will be mainly present in distantly related lineages [21]. We believe this has very limited clinical impact as resistance variants are described in relation to the H37Rv reference genome and any two *Mtb* strains have a nucleotide similarity of ≥99.8% [1]. Changing the reference genome in MAGMA would be a complex process as all tools requiring the reference genome would need to be retested.

## Conclusion

MAGMA is novel bioinformatics pipeline that can be deployed on a wide range of computing systems for the analysis of clinical *Mtb* samples. MAGMA provides users with a wide range of automated, standardized, reproducible data and visual outputs that can be used for public health and to guide the treatment of drug resistant tuberculosis.

## Methods

### Computational requirements and flexibility

The MAGMA pipeline is implemented in the Nextflow workflow management tool [22] and can be used on multiple platforms including a personal Linux computer, a shared institutional server, a cluster system such as the open source SLURM [23], a portable batch system (PBS) [24], or a cloud batch computing platform such as Amazon Web Services (AWS), Google or Azure Batch, provided that the analysis platform meets the baseline hardware requirements. Ideally, MAGMA is operated on a high-performance computing system with >32 GB RAM to limit the required runtime. Portability and reproducibility are achieved by relying on user-friendly package managers such as conda or container systems such as Docker [25–27]. MAGMA is open source and available on GitHub (https://github.com/TORCH-Consortium/MAGMA).

### Key design choices for WGS analysis by MAGMA

Four criteria are incorporated as default parameters to filter out samples that could negatively impact downstream analysis: 10x median coverage, coverage breadth below 90%, presence of mixed infections and NTM frequency exceeding 20% (Table 4). These parameters are specified in the configuration file and can be modified if required for specific research analyses.

**Table 4:** Selection Criteria to filter samples for the cohort workflow.

| MAGMA Selection Criteria | Threshold | Reasoning |
| --- | --- | --- |
| Median Coverage Depth | >10x | Sufficient coverage must be present to confidently call variants |
| Coverage Breadth | >90% | Missing data can lead to phylogenetic bias |
| Major strain frequency | >80% | Multiple infection leads to the construction of hybrid *Mtb* strains |
| NTM frequency | <20% | NTM sequences are highly similar to *Mtb* and can lead to incorrect genotyping at high contamination levels |

To allow analysis of samples with low mycobacterial burden but maintain accurate identification of genuine variants, a 10x coverage threshold was selected. This threshold ensures that almost the entire (99.995%) genome has at least one read mapped and the vast majority (97%) of genome positions have at least 5 reads associated with the reference genome position. These assumptions are based on the Poisson distribution across the genome of reads generated by sequencing platforms [28]. If the average depth of coverage is 10x, the Poisson distribution predicts that 97% of genome positions will have at least 5x coverage and 99.995% of the genome will have a coverage of at least 1x.

Because missing data in samples with low coverage breadth can lead to biased phylogenetic analyses, the MAGMA pipeline discards samples with a coverage breadth <90% [29]. Even though most samples achieve a coverage of the reference genome of 98% or greater, the threshold was relaxed to 90% to enable analysis of *Mtb* genomes with large structural deletions, which occur occasionally and can be resistance conferring [30].

While patients with tuberculosis may have mixed *Mtb* infections [31], MAGMA excludes the analysis of mixed infections when none of the strains dominate (i.e., when none of the strains have a frequency of more than 80%) because mixed infection without a dominant strain creates a hybrid strain. Hybrid strains cause long branch attraction in the phylogeny which impacts the XBS variant calling core of the MAGMA pipeline and complicates the interpretation of phylogenetic trees [32]. The threshold of a majority strain at frequency ≥80% was chosen to avoid calling false positive variants in the minority strain, which can then only be present at maximum 20%. Under a Poisson distribution, variants in the minority *Mtb* strain occurring at 20% frequency only have a 0.8% chance of being detected as a major allele (i.e., with a frequency >50%). To check the 80% threshold, we use the lineage annotation of TBProfiler [33] at the most detailed (i.e., smallest sub-) lineage level.

Patients with tuberculosis can also have mixed infection with *Mtb* and non-tuberculous mycobacteria (NTM). Reads generated from NTM genomes map to the H37Rv *Mtb* reference genome due to their high levels of similarity [6]. This complicates variant calling and phylogenetic tree construction due to the false positive detection of variants that are present in NTMs but not in the *Mtb* strain. Similar to the criterium of mixed *Mtb* infection, the MAGMA pipeline discards samples from downstream workflows when the frequency of NTMs is >20%.

### Pipeline architecture and overview of the key workflows

The MAGMA pipeline consists of multiple per-sample analysis steps followed by a cohort analysis (Figure 5). Raw sequencing files are first quality-controlled and mapped to the H37Rv *Mtb* reference genome. Variant calling is then performed for major, minor, and structural variants for all samples. Samples that pass the quality control are grouped in the cohort analysis after which they are genotyped, and SNP variants are recalibrated and filtered. Filtered SNPs and unfiltered INDELs are then used to determine the resistance profile of individual samples, the presence of potential transmission clusters and to construct a phylogenetic tree of the sample dataset.

**Figure 5:**
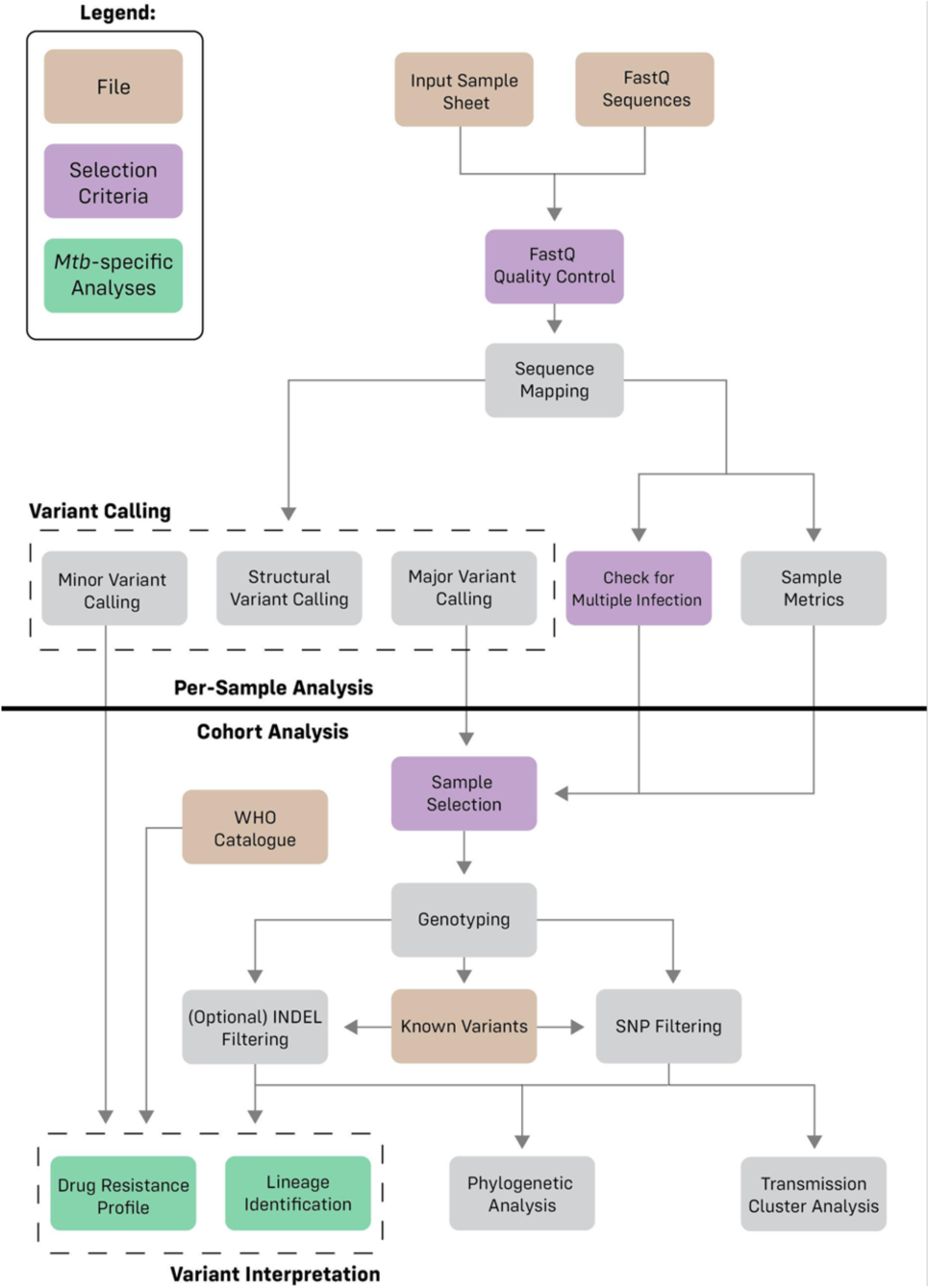
Diagram representing the different analyses which occur in the pipeline. The pipeline in split into two stages: per-sample analysis and cohort analysis

### Input files

MAGMA uses the raw FASTQ paired-end sequence output from an Illumina sequencing platform. The MAGMA pipeline is not yet validated for handling sequencing data from other platforms such as PacBio and Oxford Nanopore Technologies. MAGMA also requires a sample sheet with information on library, flowcell, and index sequence for each sample, information which is critical for the Genome Analysis Toolkit (GATK) to distinguish between genuine variants and errors introduced by sequencing.

### Quality control and mapping workflow

A visual overview of the quality control (QC) and mapping workflow is provided in supplementary figure 5. The MAGMA pipeline starts the individual sample workflow by checking the FASTQ files. FastQC is used to generate QC metrics for the sequence libraries [34] and reports are collated by MultiQC [35]. These can be used to investigate the presence of excessive non-*Mtb* sequences (as indicated by the GC%) or suboptimal sequencing quality as (indicated by low base quality).

Next, the Burrows-Wheeler Aligner (BWA) software package is used to map the sequence reads to the H37Rv reference genome (NC_000962.3) using the Maximal Exact Match (MEM) algorithm. This BWA-MEM approach was selected for its fast and reliable mapping [36]. The local alignment implemented in BWA-MEM and subsequent soft-clipping trims sequencing read ends for adapters and low-quality bases. The MAGMA pipeline uses the default BWA-MEM parameters except for increasing the seed length from 20 to 100bp. This longer seed length was implemented because it speeds up the alignment and greatly minimizes the level of contamination, including close related contaminants such as NTMs. The resulting BAM files are then deduplicated using GATK, which recognizes the presence of multiple libraries of the same sample using the information in the sample sheet. SAMtools stats and flag stats collect metrics on the quality of the library, sequence reads and the assembly [37], which are collected in the summary statistics files. GATK’s CollectWgsMetrics is used to collect metrics on depth and breadth of genomic coverage [13], which are output to a summary stats QC file. The QC steps ensure that only samples with a breadth of coverage (1x) ≥90% and a median coverage ≥10x proceed to the Cohort Workflow.

Next, LoFreq [38] is used to detect the presence of NTMs by inferring the frequency of variants at position 1472307 in the H37Rv reference genome [39]. For the assessment of the frequency of NTM reads, only well-mapped reads (i.e., mapping quality (MQ) = 60) with high-quality bases (i.e., base quality (BQ) >20) are considered. The output of the LoFreq analysis is also used as input for TBProfiler to exclude samples with mixed infection where there is no dominant strain.

### Sample level variant calling workflow

Supplementary figure 6 gives a visual overview of the variant calling workflow which starts with an optional GATK Base Quality Score Recalibration (BQSR) step to recalibrate the base qualities scores of likely systematic sequencing errors. Thereafter, the workflow branches into three sub-workflows: major variant calling, minor variant calling and structural variant calling.

For major variant calling, MAGMA uses the GATK HaplotypeCaller tool to identify “active” regions in the genome where close potential variants exist. GATK considers the ploidy of one for *Mtb* and realigns the reads in the “active” regions. The pipeline then calls likely haplotypes in the form of single nucleotide polymorphisms (SNPs) or insertions and deletions (INDELs). The resulting variants are exported to an intermediate ‘gVCF’ file for downstream joint variant calling and filtering.

Minor variant calling relies on LoFreq [38, 40]. In MAGMA, LoFreq calls only minor variants with a perfect mapping quality (MQ=60) to reduce the false positive rate. The LoFreq indel quality command is first run to assign quality scores to INDELs. SNPs and INDELs are then called simultaneously and reported in a ‘LoFreq VCF’ file. There is no default minimum threshold to ensure maximum sensitivity for the detection of minor variants. Because minor variants may only be of clinical relevance above a certain allele frequency, an option is provided to filter the VCF based on a specified threshold. The LoFreq VCF file is later used for resistance analysis but not passed through the XBS variant calling core.

MAGMA uses Delly [41] to identify large structural variants (SVs). Delly was designed for the analysis of diploid genomes and this incorrect ploidy is used advantageously in MAGMA to reduce the false positivity rate. In cases where Delly interprets a SV as heterozygous (i.e., where the SV and reference allele exist at the same position) they are assumed to be the result of contaminant sequences and are discarded from the analysis. SVs interpreted by Delly to be homozygous alleles are assumed to be potential SV variants and are exported to a ‘Delly VCF’ file. A report is created that lists SVs occurring in the WHO Tier 1 and 2 candidate drug resistance genes [42]. This information can be used to manually assess the relation of the presence of an SVs and drug resistance. The Delly VCF file is also not passed through the XBS variant calling core.

### Cohort level variant calling workflow

A visual representation of the cohort workflow is shown in supplementary figure 7. Unique to MAGMA is that variant calling is done for the entire dataset rather than on a sample-by-sample basis. Performing a cohort analysis allows GATK to confidently identify variants even if they occur at very low depth of genomic coverage (between 5x and 20x coverage) in some of the samples included in the analysis. The cohort workflow uses the multi-sample gVCF file, which combines the gVCF files of all samples that pass QC. A sample metrics summary file allows the user to investigate why certain samples were excluded from the cohort analyses.

The next step within the cohort workflow is genotyping. In *Mtb*, the majority allele is assigned as the genotype and used for phylogenetic and transmission cluster analyses. Genotyped variants are annotated using SnpEff [43], which predicts mutational effects using the pre-built ‘Mycobacterium_tuberculosis_h37rv’ reference. To better reflect the clinically relevant promoter region size in *Mtb*, the default up- and downstream interval size of 5000bp in SnpEff was reduced to 100 bp [44]. SnpEff allows a single variant to have multiple annotations, with each annotation added in the form of an ANN field [45] to the INFO column in the multi-sample VCF file. When two variants occur in the same codon, SnpEff annotates these variants as two distinct SNPs, because both variants may not occur together in all samples represented in the cohort VCF file. SnpEff therefore annotates SNPs in the same codon separately instead of as Multiple Nucleotide Polymorphisms (MNPs) which could impact the predicted effect of the variant. In MAGMA, each sample is therefore processed and annotated separately during resistance calling to avoid potential mis-annotation of MNPs for drug resistance calling.

Next, genotyped SNPs are filtered using GATK Variant Quality Score Recalibration (VQSR). This is done in an allele-specific fashion to distinguish alleles originating from contaminating sequences from alleles that occur in the *Mtb* genome. VQSR uses a truth set of high confidence genetic variants. For *Mtb*, the truth set was created as a combination of lineage markers [18] and high confidence drug resistance markers [46] and variants in rRNA genes are excluded because they have unusual variation in terms of statistical annotations [6]. VQSR starts by identifying “truth set” variants and examines their statistical annotations. Two models are built: a ‘true model’ and a ‘false model’ for the worst performing variants. All variants in the data set are assigned a variant quality score log-odds ratio (VQSLOD) based on their likelihood of falling in the true model rather than the false model. The VQSLOD cut-off score is determined by selecting a threshold that ensures 0.999 sensitivity for the truth set. This high sensitivity guarantees that nearly all true variants are identified at the cost of a very low number of false positive variants. While some pipelines exclude statistical annotations that do not show sufficient variation to distinguish true variants from false variants [13], MAGMA uses all statistical annotations and excludes the least informative annotation in successive rounds. The best-performing filtering run is then selected based on the 0.999 VQSLOD scores threshold.

Finally, GATK VQSR is used to filter INDELs by means of the same progressively minimal model building protocol implemented for SNP filtering. While the AS_MQ parameter is generally excluded for INDEL calling, MAGMA includes the AS_MQ parameter because it is essential for distinguishing true variants from contaminants, even though this means that MAGMA may miss large INDELs with low mapping quality.

Because VQSR performance improves if a high number of “truth set” variants are present in the multi-sample VCF file, the MAGMA pipeline is optimally run with a data set of at least 30 samples that are not clonally related and contain a diversity of lineage and/or DR markers. The MAGMA pipeline provides the possibility to include a dataset of 334 samples from the South African EXIT-RIF study [47]. Users can also opt to upload a dataset from their own region if it contains at least 30 non-clonally related samples.

### Drug resistance analysis

The MAGMA pipeline outputs two separate sets of resistance calls for variants in candidate drug resistance genes. One uses GATK-filtered SNPs and unfiltered INDELs to identify major (i.e., present at ≥50%) variants. The other set reports minor variants identified by LoFreq to identify the presence of heteroresistance. For each set, MAGMA implements TBProfiler *v5.0* to determine the drug-resistance profile of a sample [20, 33]. TBProfiler *v5.0* reports all variants associated with (interim) resistance as listed in the WHO catalogue as well as all variants in the tier 1 & 2 genes candidate resistance genes for which the association with resistance is unknown and variants not listed in the WHO catalogue [42]. All outputs are combined in a per sample excel sheet.

### Phylogenetic and cluster analysis

For the phylogenetic and cluster analyses, filtered SNPs are concatenated into a multi-FASTA file. For phylogenetic analyses, sites of the *Mtb* genome represented by <95% of samples, common drug resistance variants [46], and variants in rRNA genes are excluded from the multi-FASTA file. Variants present in <95% of samples are excluded because including these sites can introduce phylogenetic bias. Variants in drug-resistance conferring genes are excluded due to their positive selection, which can result in phylogenetic biases [48]. rRNA gene variants are excluded as they occasionally represent false positive variants [6]. The multi-FASTA file is then used to calculate SNP distances between all sample pairs [49], once excluding and once including the complex regions of the *Mtb* genome [50]. The two most frequently used SNP cut-offs (5bp and 12bp) are used as the cut-off for cluster identification with ClusterPicker [51].

IQtree [52] is used to identify the most likely substitution model and to compute the Maximum Likelihood tree. To save computing time, bootstrapping is not performed in the default setting, but users can opt for an ultra-fast or regular bootstrap option [53]. The resulting tree can be visualized with iTOL, which can annotate phylogenetic trees with drug-resistance and lineage markers using the annotation files as created by TBProfiler [33, 54]. Phylogeny is inferred twice, once including and once excluding complex regions. MAGMA also performs cluster analysis based on the calculated SNP distances. Clusters identified by ClusterPicker are visualized using FigTree [55].

### Comparison of MAGMA and MTBseq pipelines

To compare the performance MAGMA and MTBseq pipelines and the interpretation of the results generated, we analyzed positive MGIT samples collected from 79 consecutive participants in the SMARTT clinical trial. The FASTQ files were run through MAGMA v.1.0.1 (https://github.com/TORCH-Consortium/MAGMA/) and MTBSeq v1.1. (https://github.com/ngs-fzb/MTBseq_source) using default parameters. For drug resistance analyses, variants annotated by one or both pipeline and their relevance regarding drug resistance to antituberculosis drugs as generated by the two pipelines were compared. The comparison was done in detail for one patient (TB280), summarized for all 79 samples, and differences in resistance calling were investigated. Because MAGMA and MTBseq sometimes output different annotations for identical variants, the script verified the genome position of the reported nucleotide change.

For phylogenetic analyses, the same Maximum Likelihood phylogenetic inference was performed for both pipelines. Given that MTBseq by default produces a FASTA file without complex regions for phylogenetic analyses, the comparison of phylogenetic results excluded information on complex regions for both pipelines. A second phylogenetic output was generated for MTBseq by excluding samples with a depth of coverage below 30x or a breadth of coverage below 95%, as suggested in the MTBseq frequently asked questions document. To compare the phylogenetic outputs, heatmaps were generated using the python packages seaborn (v.0.12.2) and matplotlib (v.3.8.0) from the pairwise SNP distances calculated for the phylogenies’ multi-FASTA files and co-phylogenetic face-to-face trees were plotted using the R packages ape (v.5.7-1) and phytools (v.1.9-16).

## Declarations

### Ethics approval and consent to participate

The research was performed in accordance with the Declaration of Helsinki. Collection of the clinical strains and the determination of the genotypic drug resistance profile was approved by appropriate ethics committees. For the analysis of the four samples from the SMARTT study, ethics approval was obtained in context of the SMARTT trial from ethics committees of the University of the Free State Health Sciences Research Ethics Committee (UFS-HSD2019/0364/2004), Stellenbosch University Health Research Ethics Committee (N19/07/100), and the University of Antwerp (21/18/239).

### Consent for publication

Not applicable

### Availability of data and materials

The pipeline is available from https://github.com/TORCH-Consortium/MAGMA/ and the authors confirm all supporting data, code and protocols have been provided within the article or through supplementary data files. FastQ files for the 79 samples are accessible in the ENA.

### Competing interests

A.S. was an employee of Seqera Labs S.L. during the initial development of the pipeline. Seqera Labs S.L. is the driving company behind the open source Nextflow software.

A.S. has also worked on a Nextflow wrapper for the MTBseq pipeline. The remaining authors declare that they have no competing interests.

### Funding

This research was funded by the Research Foundation Flanders (FWO) strategic basic research grant 1SB4519N, (PhD funding for L.V.), the FWO Odysseus grant G0F8316N, and the FWO applied biomedical research with a primary social finality T001018N. The funding body played no role in the design of the study and will not play any role in data collection, analysis, and interpretation of data and in writing the manuscript.

### Authors’ contributions

Authors T.H.H, L.V., and A.S are equal contributors

T.H.H.: Conceptualization, design of the work, analysis, interpretation of data, creation of new software, writing sections of the first draft of the manuscript, and critically reviewing the full manuscript

L.V.: Conceptualization, design of the work, analysis, interpretation of data, creation of new software, writing sections of the first draft of the manuscript, and critically reviewing the full manuscript

A.S.: Analysis, interpretation of data, creation of new software, writing sections of the first draft of the manuscript, and critically reviewing the full manuscript

V.R.: Analysis, interpretation of data, writing sections of the first draft of the manuscript, and critically reviewing the full manuscript M.d.D.F.: Analysis and interpretation of comparisons between pipelines.

R.M.W.: Conceptualization, generating clinical sequencing data, critically reviewing the full manuscript

A.VR.: Conceptualization, design of the work, acquisition of funding, collection of clinical sequencing data, writing sections of the first draft the manuscript, and critically reviewing the full manuscript

## Data Availability

All data produced in the present work are contained in the manuscript, or available upon reasonable request to the authors.

## Acknowledgements

We would like to thank Jody Phelan for continued support with TBProfiler, the SMARTT trial participants whose data is included in the manuscript, and the SMARTT team (Gavin Churchyard, Salome Charalambous, Maraba, Felex Ndebele, Zandile Sibeko, Pulane Segwaba, S’thabiso Bohlela, Anneke Van der Spoel Van Dijk, Emmanuel Ayodeji Ogunbayo, Mhlambi Nomadlozi, Emilyn Costa Conceicao, Felicia Wells, Astrid Paulse, Fanampe Boitumelo, Trang Tu) for enrolling patients, collecting samples and performing the laboratory processes of whole genome sequencing.

## Notes

### Author Declarations

Ethics approval was obtained in context of the SMARTT trial from ethics committees of the University of the Free State Health Sciences Research Ethics Committee (UFS-HSD2019/0364/2004), Stellenbosch University Health Research Ethics Committee (N19/07/100), and the University of Antwerp (21/18/239).

