## supplementary figures for "The MAGMA pipeline for comprehensive genomic analyses of clinical *Mycobacterium tuberculosis* samples"

Supplementary Figure 1: Phylogenetic tree created by (a) unfiltered MTBseq and (b) MAGMA excluding complex regions

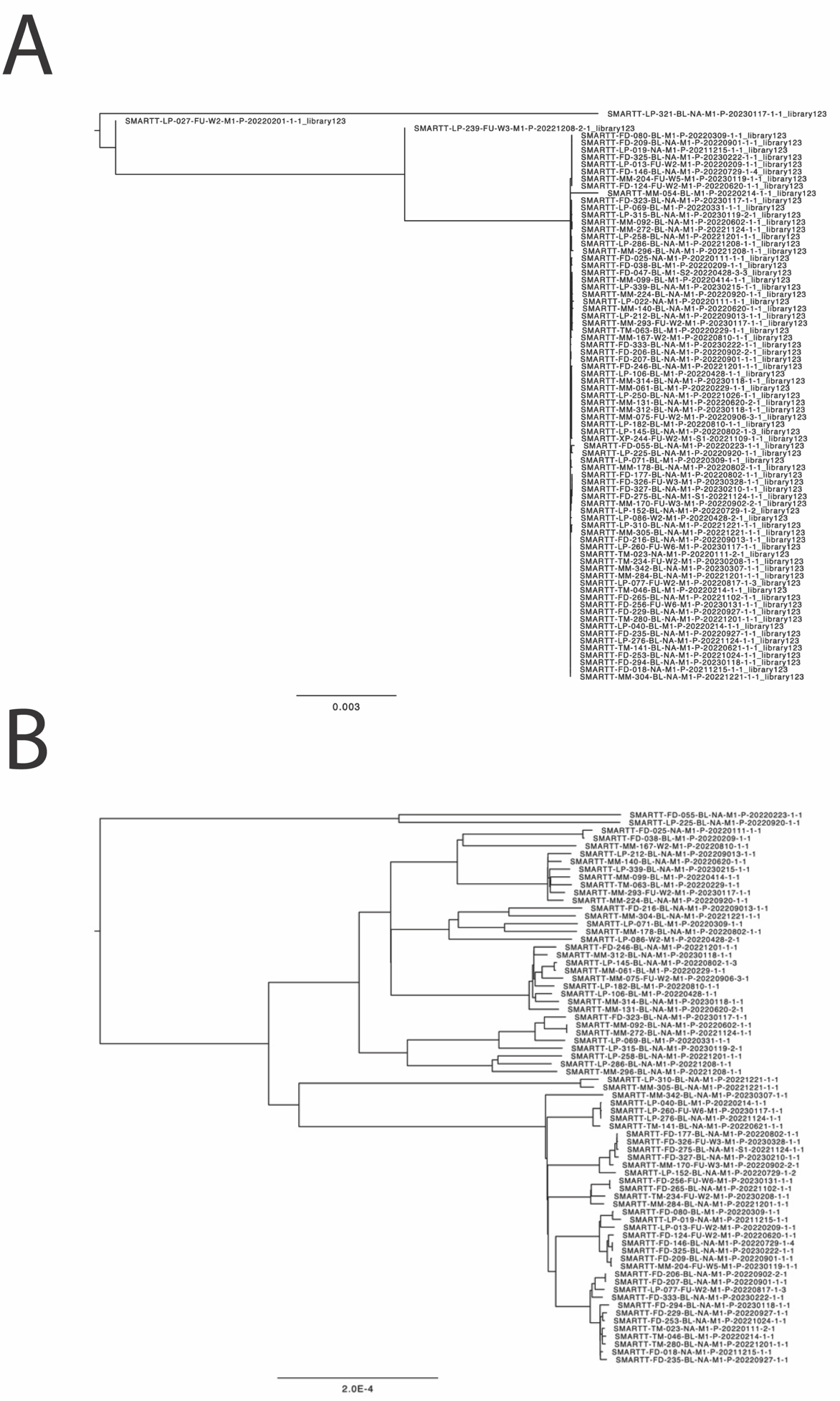

Supplementary Figure 2: Face to face comparison of the default MAGMA (right) and filtered MTBseq tree (left)

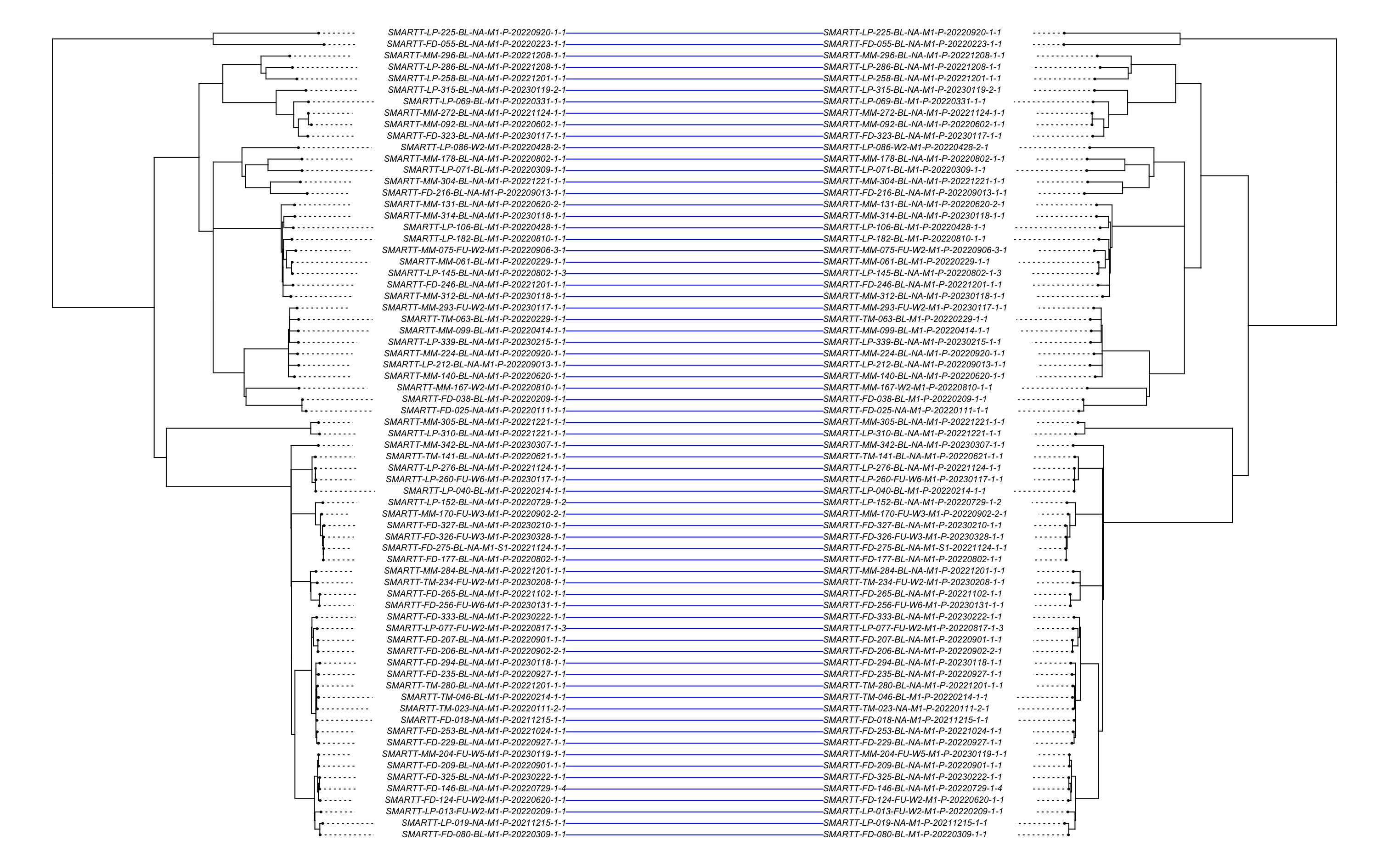

Supplementary Figure 3: Heatmap of the (a) MAGMA phylogenetic tree excluding complex region, (b) default MTBseq tree, and (c) filtered MTBseq tree

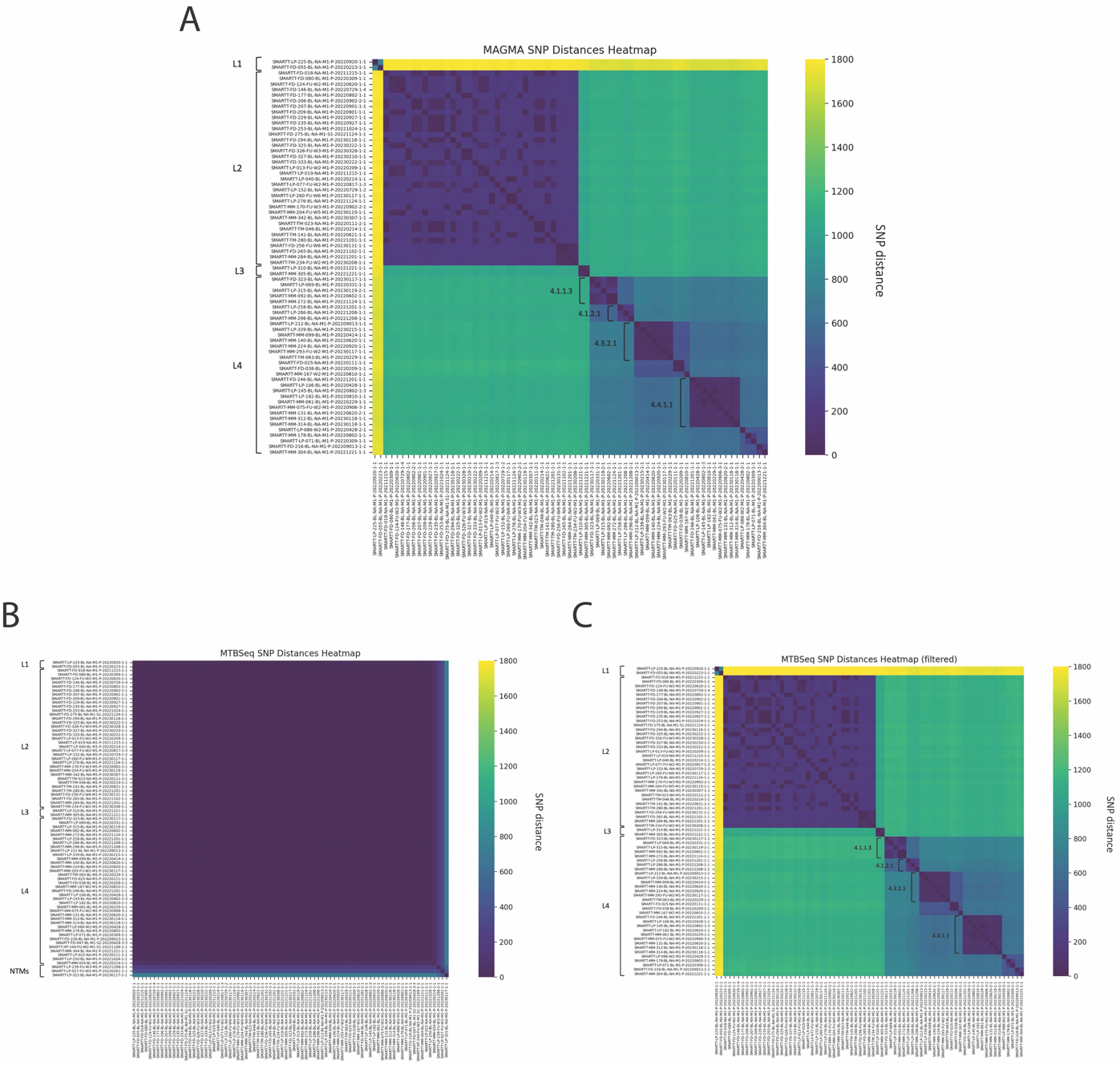

Supplementary Figure 4: Quality Control and Mapping Workflow

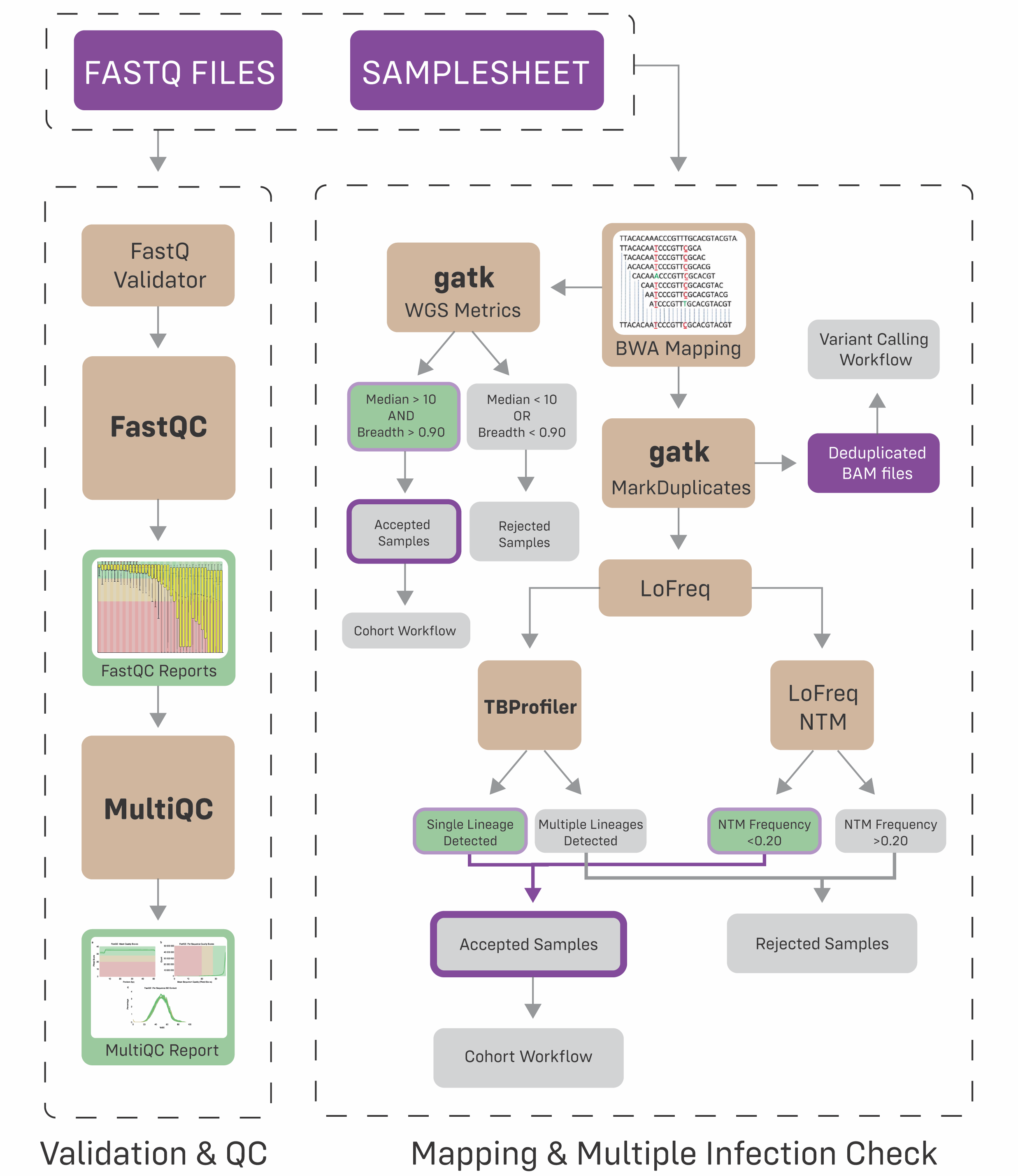

Supplementary Figure 5: Per Sample Variant Calling Workflow

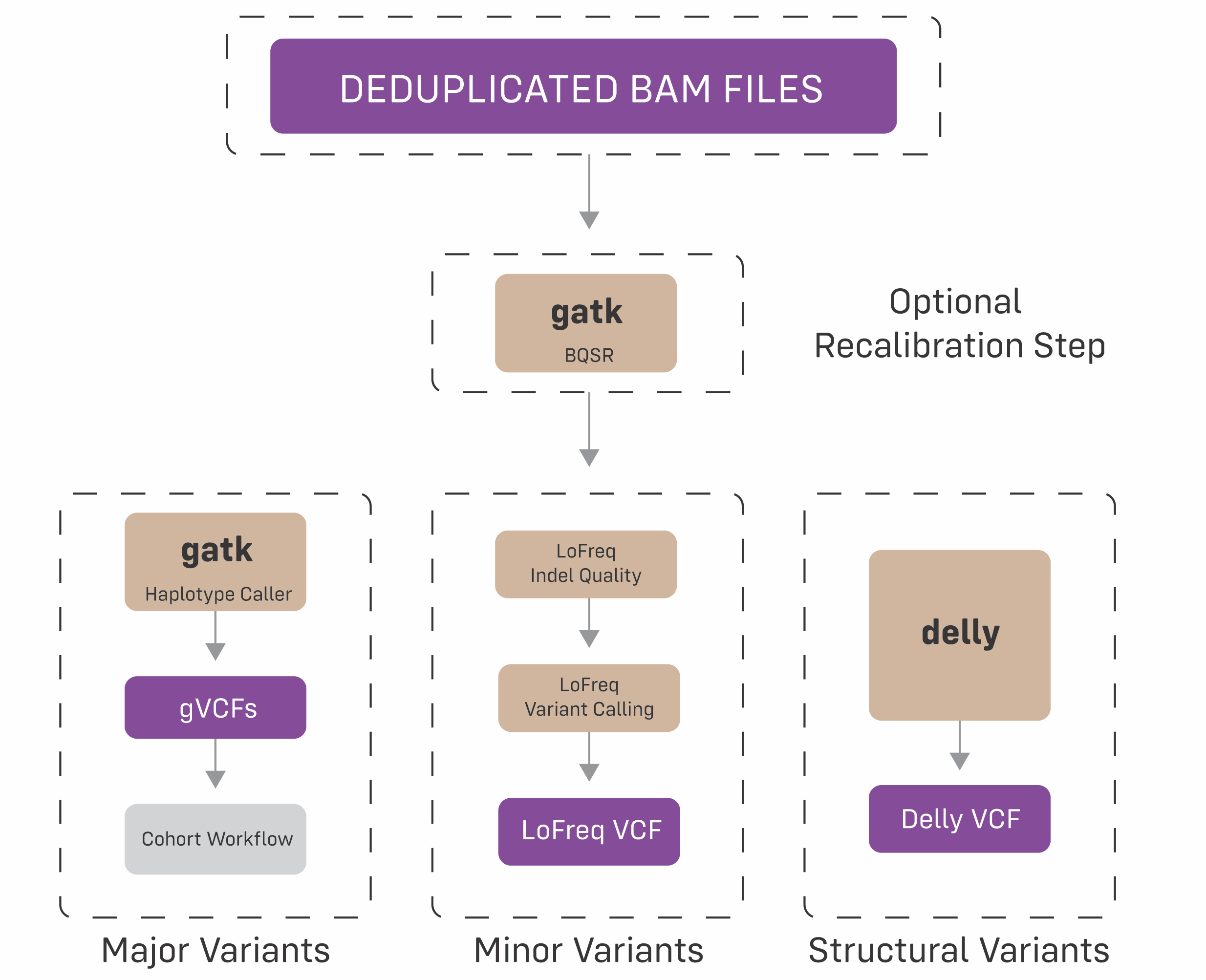

Supplementary Figure 6: Cohort Workflow

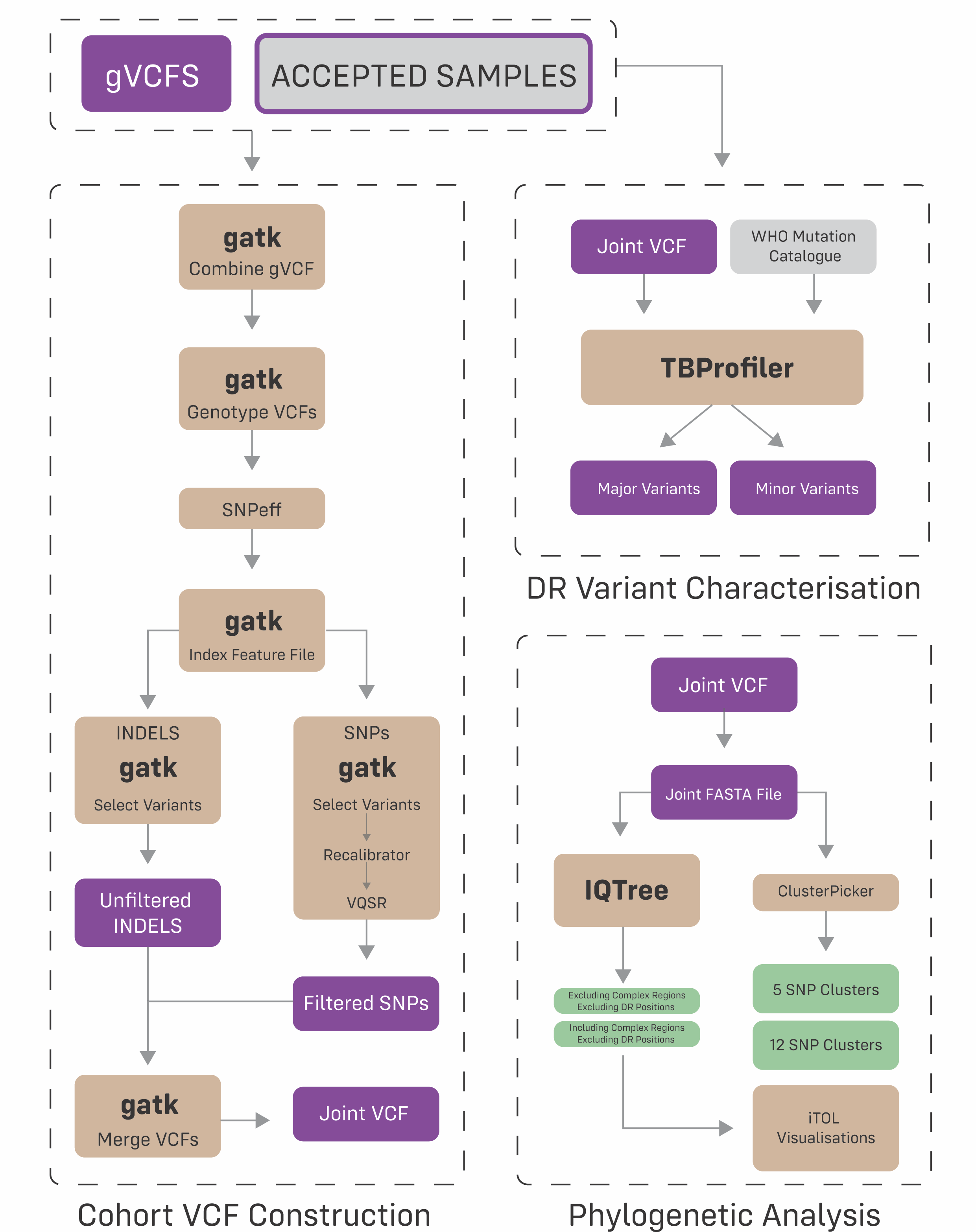
